# Bridging Brain Signals and Self-Reported Symptoms: An AI-Driven, High-Sensitivity Model for Detecting Suicidality in Major Depressive Disorder

**DOI:** 10.1101/2025.11.12.25340074

**Authors:** Chih-Chung Huang, Chun-Lun Hsu, Yi-Guang Wang, Tien-Yu Chen, Chun-Yen Chen, Ta-Chuan Yeh, Tsai-Hsun Huang, Fu-Yu Yu, Yi-Hung Liu, Che-Sheng Chu, Hsin-An Chang

## Abstract

**Background:** Major depressive disorder (MDD) with suicidality represents a significant public health concern, as suicide ranks among the leading causes of death worldwide. While electroencephalography (EEG) has shown promise in depression diagnosis, its utility in identifying suicidal risk remains underexplored. This study aims to develop and validate a Suicidal Risk Index (SR Index) using EEG biomarkers and machine learning (ML) algorithms for distinguishing between MDD patients with and without suicidality.

**Methods:** In this retrospective observational study, resting-state EEG data were collected using Stress EEG Assessment (SEA) system. SR Index was developed by integrating the PHQ-9 scale with EEG-derived features, including band power, coherence, and fractal dimension, optimized using ML algorithms to enhance accuracy.

**Results:** The study included 268 participants (159 without suicidality, 109 with suicidality). The SR Index demonstrated robust discriminative ability with an AUC of 0.8117 (*p*=2.63×10^-18^). At the optimal cutoff value of 8, the model achieved 88.99% sensitivity, 57.23% specificity, 67.86% positive predictive value, and 78.85% negative predictive value, with a balanced accuracy of 73.11%.

**Conclusions:** The SR Index shows promise as an objective tool for identifying suicidality in MDD patients, potentially complementing traditional clinical assessments. This approach may enhance early detection and risk stratification in clinical settings, potentially improving suicide prevention strategies.

**Highlights:**

- Novel EEG- and ML-based Suicidal Risk Index distinguishes MDD patients with and without suicidality.
- SR Index achieves 88.99% sensitivity and 73.11% balanced accuracy in identifying suicidal risk.

## 1. Introduction

Major depressive disorder (MDD) is a prevalent and disabling mental health condition characterized by persistent depressed mood, diminished interest or pleasure in daily activities, changes in appetite, sleep disturbances, psychomotor alterations, fatigue or loss of energy, feelings of worthlessness or excessive guilt, diminished ability to concentrate or indecisiveness, hopelessness, and suicidal ideation or attempts lasting for at least two consecutive weeks[1]. The global point prevalence of MDD is approximately 4.7% [2], with a twelve-month prevalence of 7% in the United States [1], 6.38% in Europe [3], and a lifetime prevalence of 3% in Japan[2]. This condition affects around 320 million people worldwide [4, 5]. One of the most consistent findings in MDD epidemiology is a higher prevalence in females, with rates about twice as high as in males, peaking in adolescence and then stabilizing [1]. MDD is a leading cause of disability worldwide and significantly impacts the quality of life and overall functioning of affected individuals [5, 6].

Of particular concern is the elevated risk of suicidal ideation and behaviors in MDD patients, with a suicide risk rate of approximately 10-15% during major depressive episodes [2]. According to the World Health Organization, suicide is a leading cause of death globally, with 703,000 deaths by suicide in 2019. The global age-standardized suicide rate is 9.0 per 100,000 population, with higher rates in men (12.6 per 100,000) compared to women (5.4 per 100,000)[7]. Each year, approximately 160 million people experience suicidal ideation, with around 16 million suicide attempts, and among those who attempt suicide, 1.6% will commit suicide within the next 12 months [8]. Suicidal ideation can range from passive thoughts, such as a desire not to wake up in the morning, to more serious intentions involving specific plans and preparations [1]. Therefore, identifying suicidal ideation is critical for effective prevention.

To date, most research on suicide has focused on identifying risk factors and promoting strategies to reduce overall suicide rates in specific populations. The most consistently described risk factor is a prior history of suicide attempts or threats. Anhedonia, or the inability to experience pleasure, is strongly associated with suicidal ideation [1]. Other features linked to increased suicide risk include being single, living alone, social isolation, early life adversity, access to lethal means such as firearms, sleep disturbances, cognitive and decision-making impairments, prominent feelings of hopelessness, stressful life events, chronic medical conditions, financial distress, and substance abuse[1, 8]. Identifying and differentiating those at heightened risk of suicide within the broader MDD population is therefore critical for improving treatment outcomes and implementing preventive interventions.

Despite advances in the diagnosis and management of MDD, reliably identifying patients with suicidal ideation or behaviors remains challenging due to the heterogeneity of its etiology and varied psychopathological manifestations[9]. Depression and suicide lack clear biomarkers or gold standards for diagnosis, and they are primarily diagnosed based on clinical symptoms. Rating scales such as the Patient Health Questionnaire-9 (PHQ-9) and the Beck Depression Inventory are commonly used to screen for depressive symptoms and suicide risk [8, 10, 11]. However, these tools have limitations due to their reliance on self-reporting, lack of balance between sensitivity and specificity in detecting acute suicidal tendencies, and the potential for increased clinical workload due to false-positive cases requiring psychiatric assessment or hospitalization [8]. Moreover, patients’ willingness to share information can affect the accuracy of these assessments. Although the Columbia Suicide Severity Rating Scale offers a more effective alternative compared to traditional clinical assessment tools, it is impractical to screen every patient at each clinical encounter [12]. Thus, there is a pressing need for objective biomarkers that can predict suicide risk within an actionable time frame.

Many studies have attempted to apply machine learning (ML) techniques to classify resting-state EEG signals in MDD patients. However, the accuracy of classification based on EEG and ML varies widely across studies, ranging from about 60% to 99%, with most studies having small sample sizes (fewer than 100 subjects) and using low-density EEG signals with only a few electrodes.

Additionally, the lack of diverse data hinders the generalizability of these algorithms or models. A prior study utilizing ML and large-scale, multi-center EEG data achieved 84% accuracy in individual classification, demonstrating the effectiveness and applicability of EEG- and ML-based computer-aided diagnosis (CADx) in MDD detection[4]. However, no such studies have focused on CADx for MDD patients at risk of suicide.

In this study, we aim to compare the neurophysiological profiles of MDD patients with and without suicidality using EEG- and ML-based biomarkers. By integrating EEG data with clinical assessments, including the PHQ-9 scale, and leveraging ML algorithms, we seek to establish more accurate indicators of suicide risk, which we term the Suicidal Risk Index (SR Index). The primary objective of this study is to elucidate the differences in brainwave patterns between these two groups, contributing to the development of more precise diagnostic tools and targeted therapeutic interventions for MDD patients at risk of suicide.

## 2. Materials and Methods

### 2.1. Study Design, Participants and Ethical Approval

This study employed a retrospective observational design conducted at the Tri-Service General Hospital (TSGH), a tertiary medical center in Taiwan. The study protocol received approval from the Institutional Review Board (IRB) of TSGH (IRB# A202305083).

The study population comprised patients diagnosed with MDD. Inclusion criteria were as follows: (1) age 20 years or older; and (2) MDD diagnosis confirmed by two board-certified psychiatrists based on the criteria outlined in the Diagnostic and Statistical Manual of Mental Disorders, Fifth Edition (DSM-5) [1]. Exclusion criteria encompassed: diagnosis of other psychiatric disorders (including psychotic disorders, bipolar disorder, obsessive-compulsive disorder, autism spectrum disorder, neurodevelopmental disorders, substance use disorders [excluding caffeine and/or tobacco], and neurocognitive disorders); diagnosis of organic brain disorders (such as stroke, epilepsy, brain tumors, arteriovenous malformations, or meningitis); and use of anticonvulsants (e.g., clonazepam, diazepam) within seven days prior to assessment.

Enrolled participants underwent assessment for suicide risk using the item 3 of 17-item Hamilton Depression Rating Scale (HDRS-17). Patients scoring 1-4 on this item were classified into the MDD with suicidality group, while those scoring 0 were assigned to the MDD without suicidality group [13].

Each participant completed the PHQ-9 [10] and underwent electroencephalogram (EEG) testing using the Stress EEG Assessment (SEA) system developed by HippoScreen Neurotech Corp.. The SEA system, developed in collaboration with four major medical centers in Taiwan, utilizes EEG data collected from patients with depression [4]. The system incorporates a quantitative index, the SEA Index, which employs ML techniques to nonlinearly transform the feature space of EEG data, classify the data, and subsequently discretize and stratify it to generate a quantitative index ranging from 1 to 10. In 2023, the SEA Index received approval from the Taiwan Food and Drug Administration (TFDA) as an AI-based Software as a Medical Device (SaMD). It serves as a reference tool for assisting in the diagnosis of suspected depression, with higher SEA Index scores indicating a greater likelihood that the EEG data is associated with depressive symptoms.

### 2.2. Apparatus

The SEA system utilized a 24-inch flat monitor for stimulus presentation, with participants seated at a distance of 70 cm from the screen. EEG data were recorded using an 8-channel WaveGuard EEG cap (ANT Neuro, Netherlands) connected to an HNC amplifier (HippoScreen Neurotech Corp.). The amplifier was set with a gain of 50, 24-bit A/D resolution, and a 500 Hz sampling rate. The sintered ring electrodes used were made of aluminum/silver chloride (Al/AgCl), and the electrode montage was based on the extended international 10–20 system. Electrode gel (Gelaid, provided by Nihon Kohden) was applied to maintain impedance below 10 K Ohms. EEG signals were referenced on-line to the right mastoid (A2 electrode).

### 2.3. Resting-state EEG Acquisition, Data Collection, and Preprocessing

Participants were seated in a light- and sound-attenuated room on a comfortable chair with armrests. The WaveGuard EEG cap was positioned by aligning intersection of the nasion, inion, and preauricular points with the Cz position, ensuring proper electrode-scalp contact. Conductive gel was applied to each electrode to optimize conductivity. Once the cap was properly in place, EEG data collection commenced.

Ninety seconds of eyes-open resting-state EEG signals were recorded following a 5-second prompt signal. Participants fixated on a crosshair (black cross on gray background) displayed 70 cm away, maintaining open eyes and minimizing any head or body movement while remaining alert.

During preprocessing, the vertex electrode (Cz) served as the reference signal. A 2–60 Hz bandpass filter was applied digitally. The 90-second resting-state EEG data from seven electrodes were segmented into 15 non-overlapping 6-second epochs for subsequent analysis.

### 2.4. Processing of The Stress EEG Assessment (SEA) System and SEA Index

EEG signal analysis involved extracting frequency domain features using band power (BP) and coherence. Continuous data were transformed using Discrete Fourier Transform to convert the original EEG signals from the time domain to the frequency domain. BP values were computed for predefined frequency bands: theta (4–8 Hz), alpha (8–13 Hz), beta (13–30 Hz), and gamma (30–45 Hz). Coherence between signals was calculated using Fast Fourier Transform, representing the degree of cooperation and synchronization between brain regions.

Fractal dimension analysis, specifically Higuchi fractal dimension (HFD) [14] and Katz fractal dimension (KFD) [15], was employed to describe signal complexity and hidden information within the temporal signal. HFD provides precise fractal dimension estimates but is more susceptible to noise interference, while KFD offers better noise suppression.

Feature selection utilized heuristic methods such as Sequential Backward Selection to identify the most discriminative feature combinations [16], effectively removing redundant features. A Support Vector Machine (SVM) classifier was employed for feature classification [17], aiming to find an optimal separating hyperplane between classes. Non-Linear SVM was used for non-linearly separable data in higher-dimensional feature spaces.

The SVM-derived decision value underwent sigmoid function transformation to obtain a probability value, *f*(*D*). This value was subsequently graded to generate a quantitative score (SEA Index) ranging from 1 to 10, facilitating interpretation of classification results [4].

### 2.5. Statistical Analysis

Demographic and clinical characteristics were performed using IBM SPSS Statistics 26.0 software (IBM SPSS Inc., Chicago, IL, USA). Comparisons between the MDD with suicidality group and the MDD without suicidality group for age, PHQ-9 scores, and SEA Index were conducted using Student’s t-test or Mann-Whitney U test, depending on sample size and data distribution. Sex distribution differences were assessed using the Chi-square test. All statistical tests were two-tailed, with significance defined as *p* < 0.05.

Binary logistic regression analysis was employed to determine the optimal model for predicting suicidal ideation in MDD patients, distinguishing between the MDD with suicidality and MDD without suicidality groups. The model’s predictive performance was evaluated using the receiver operating characteristic (ROC) curve and area under the curve (AUC). The optimal cutoff value for differentiating MDD with suicidality from MDD without suicidality was identified using the highest Youden Index (J), which maximizes the sum of sensitivity and specificity. AUC values were interpreted as follows: >0.9 excellent, 0.8–0.9 very good, 0.7–0.8 good, 0.6–0.7 sufficient, and 0.5–0.6 poor diagnostic accuracy in distinguishing MDD with suicidality from MDD without suicidality [18].

## 3. Results

The study population consisted of 268 participants divided into two groups. The MDD without suicidality group (n=159) comprised 99 females (mean age 50.17 years) and 60 males (mean age 43.91 years). The MDD with suicidality group (n=109) included 81 females (mean age 44.5 years) and 28 males (mean age 52.07 years). Demographic analysis revealed no statistically significant differences between groups regarding age or sex distribution (Table 1).

**Table 1.** Demographic and Clinical Characteristics of MDD with Suicidality and MDD without Suicidality.

|  | MDD with<br>suicidality (n=109) | MDD without<br>suicidality (n=159) | *p*-Value* | Effect Size |
| --- | --- | --- | --- | --- |
| Sex (Female/Male) | 81/28 | 99/60 | 0.0535 | 0.2576 |
| Age, years (mean ± SD) | 46.45 ± 16.22 | 47.81 ± 16.14 | 0.4962 | 0.0416 |
| PHQ-9 Score (mean ± SD) | 16.64 ± 6.42 | 10.42 ± 7.02 | < **0.0001** | 0.4170 |
| SEA Score (mean ± SD) | 6.47 ± 2.32 | 4.92 ± 2.71 | < **0.0001** | 0.2822 |

\* Significant effects were marked with bold characters ( $p < 0.05$ ).
PHQ-9 Score=Patient Health Questionnaire-9; SEA=Stress EEG Assessment.

### 3.1. Suicidal Risk Index (SR index)

A new index termed the SR Index was designed by combining PHQ-9 scores with the SEA Index. The PHQ-9 scores were categorized into five groups based on severity levels: None-minimal, mild, moderate, moderately severe, and severe [10], as follows: [0 ≤ PHQ-9 ≤ 4, 5 ≤ PHQ-9 ≤ 9, 10 ≤ PHQ-9 ≤ 14, 15 ≤ PHQ-9 ≤ 19, 20 ≤ PHQ-9 ≤ 27]. Similarly, the SEA Index was divided into five groups: [1 ≤ SEA ≤ 2, 3 ≤ SEA ≤ 4, 5 ≤ SEA ≤ 6, 7 ≤ SEA ≤ 8, 9 ≤ SEA ≤ 10].

By combining these two indices, a 5×5 matrix was constructed, with each cell corresponding to an SR Index score. As shown in Table 2, we hypothesized that as both PHQ-9 and SEA Index scores increase, the suicide risk would also correspondingly increase. Thus, the SR Index scores were arranged to increase from left to right and from top to bottom across the matrix, indicating that higher PHQ-9 and SEA Index combinations corresponded to a higher SR Index.

**Table 2.** SR Index Matrix Combining PHQ-9 and SEA Scores*.

|  | SEA Score |  |  |  |  |
| --- | --- | --- | --- | --- | --- |
|  | 1-2 | 3-4 | 5-6 | 7-8 | 9-10 |
| PHQ-9 Score<sup>†</sup> |  |  |  |  |  |
| 0-4 | 1 | 2 | 3 | 4 | 5 |
| 5-9 | 2 | 3 | 4 | 5 | 6 |
| 10-14 | 3 | 4 | 5 | 6 | 7 |
| 15-19 | 4 | 5 | 6 | 7 | 8 |
| 20-27 | 5 | 6 | 7 | 8 | 9 |

\* The SR Index, constructed by combining PHQ-9 and SEA scores, increases from left to right (SEA) and from top to bottom (PHQ-9), indicating a higher risk of suicidal ideation with elevated combinations of PHQ-9 and SEA scores.
<sup>†</sup> PHQ-9 Score Range: 0-4: Minimal depression; 5-9: Mild depression; 10-14: Moderate depression; 15-19: Moderately severe depression; 20-27: Severe depression.
PHQ-9 Score=Patient Health Questionnaire-9; SEA=Stress EEG Assessment; SR Index=Suicide Risk Index.

To validate this hypothesis, we used different SR Index cutoff points to calculate sensitivity and specificity, and plotted the ROC curve along with calculating the AUC to evaluate the efficacy of using the SR Index for assessing suicide risk (Fig. 1). Additionally, we presented the sensitivity and specificity for different cutoff points (Table 3). The resulting AUC was 0.7854, indicating a significant difference between the MDD with suicidality and MDD without suicidality groups, with a *p*-value of 1.07×10^-15^. This result suggests that the SR Index effectively distinguishes between MDD with suicidality and MDD without suicidality patients.

**Fig. 1.**
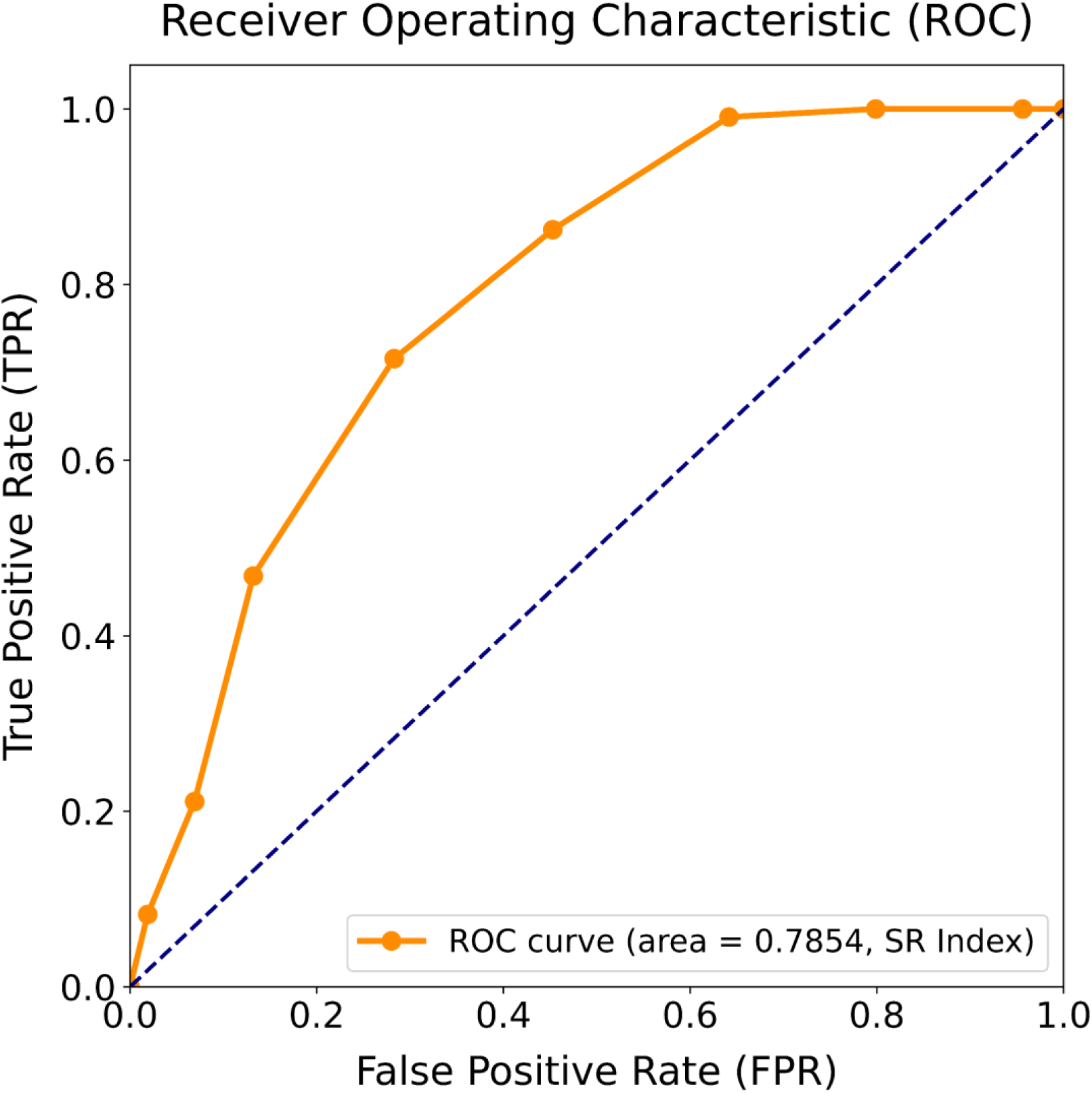
Receiver Operating Characteristic (ROC) curve analysis of the SR Index for distinguishing between MDD with suicidality and MDD without suicidality groups. The ROC curve demonstrates the diagnostic performance of the SR Index in differentiating MDD with suicidality from those MDD without suicidality. The area under the curve (AUC) of 0.7854 indicates good discriminative ability. The diagonal dashed line represents random classification (AUC = 0.5). TPR: True Positive Rate; FPR: False Positive Rate.

**Table 3.** Sensitivity and Specificity Across Different Cut-off Points for SR Index.

| Cut-off Point | Sensitivity (%) | Specificity (%) |
| --- | --- | --- |
| 1 | 100.00 | 0.00 |
| 2 | 100.00 | 4.40 |
| 3 | 100.00 | 20.13 |
| 4 | 99.08 | 35.85 |
| 5 | 86.24 | 54.72 |
| 6 | 71.56 | 71.70 |
| 7 | 46.79 | 86.79 |
| 8 | 21.10 | 93.08 |
| 9 | 8.26 | 98.11 |

SR Index=Suicide Risk Index.

### 3.2. Optimal Suicidal Risk Index (Optimal SR index)

To determine the optimal SR Index combinations from the 5×5 matrix, we employed Optuna [19], an automated hyperparameter optimization framework. Optuna uses the Tree-structured Parzen Estimator method to automatically explore different parameter combinations by defining an objective function for the optimization problem. Based on the training performance of the model, Optuna adjusts the hyperparameters iteratively, aiming to improve the efficiency of the optimization process. Specifically, we designed an objective function that generated SR Index values based on PHQ-9 and SEA Index scores (Table 4), and subsequently calculated their corresponding AUC values.

**Table 4.**
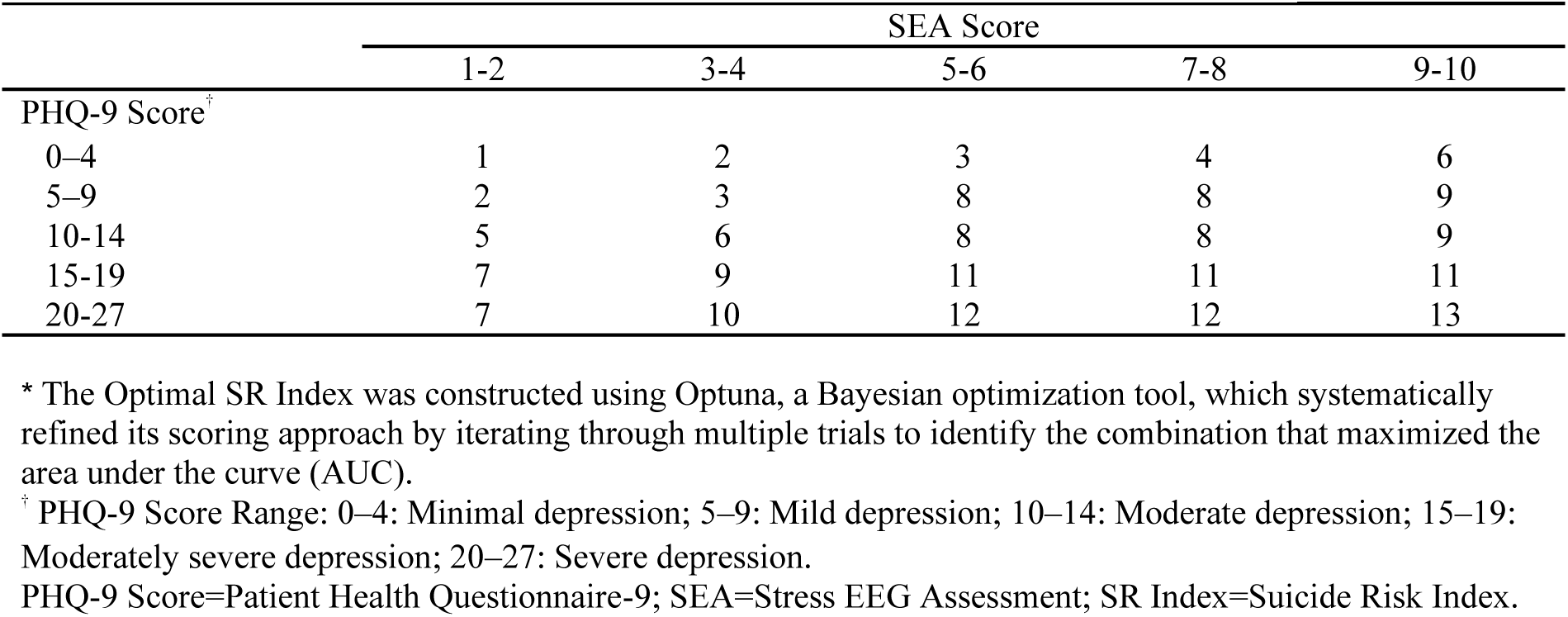
Optimal SR Index Matrix Combining PHQ-9 and SEA Scores*.

Optuna utilizes a Bayesian optimization strategy, performing multiple trials and continuously iterating to refine its approach, systematically exploring various scoring combinations. In each trial, Optuna updated its search strategy based on the results of the previous iteration to move more efficiently toward an optimal solution. Ultimately, the SR Index that maximized the AUC was identified, as shown in Fig 2. The resulting AUC was 0.8117, with a significant difference observed between the MDD with suicidality and MDD without suicidality groups (*p*-value = 2.63×10^-18^). When using a cutoff value of 8, according to Youden’s J statistic, the optimal balance between sensitivity and specificity was achieved. The values for sensitivity, specificity, positive predictive value, and negative predictive value were 88.99%, 57.23%, 67.86%, and 78.85%, respectively, with a balanced accuracy of 73.11% (Table 5).

**Fig. 2.**
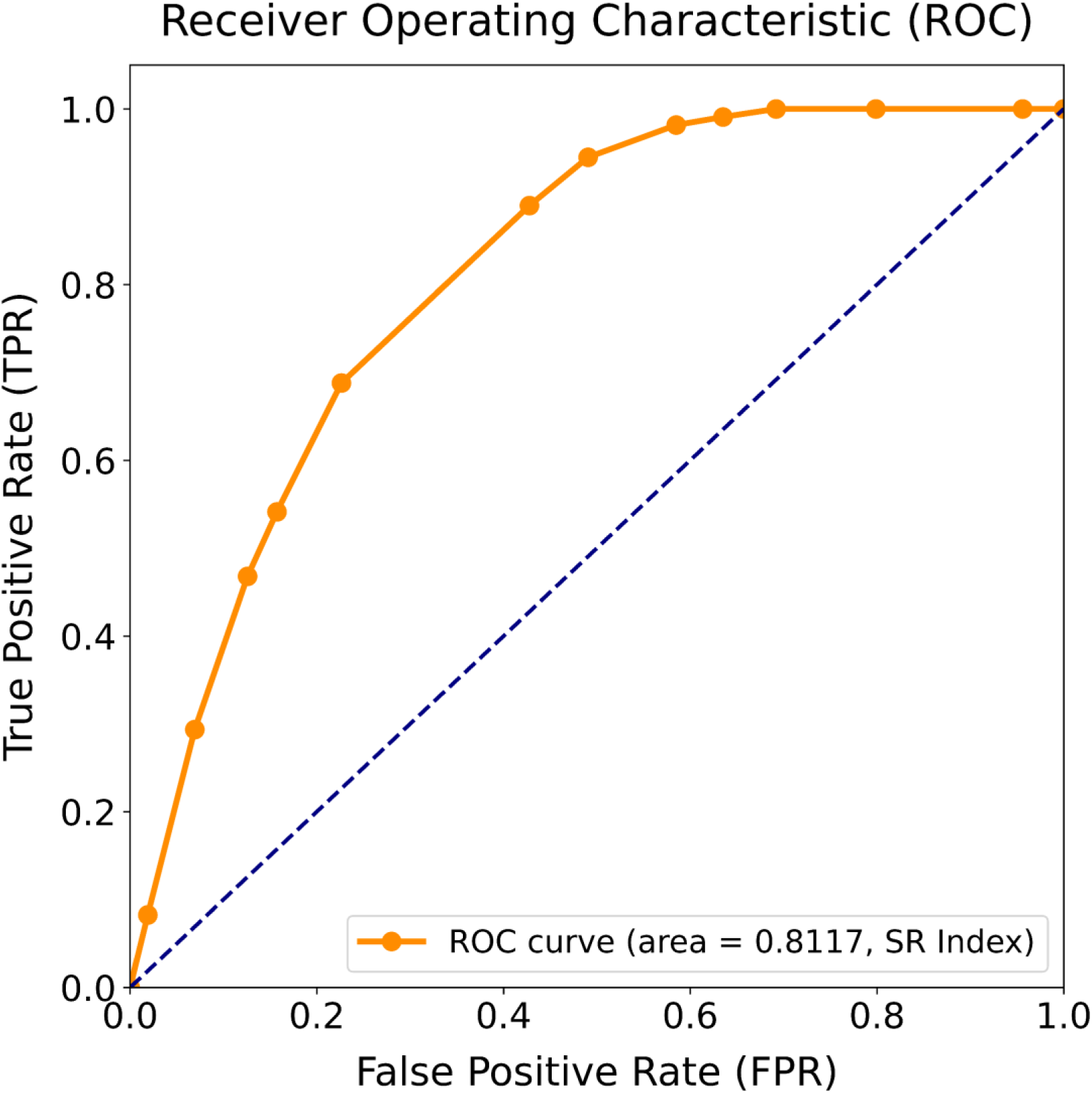
Receiver Operating Characteristic (ROC) curve analysis of the optimal SR Index for discriminating between MDD with suicidality and MDD without suicidality groups. The ROC curve demonstrates the diagnostic performance of the SR Index in differentiating MDD with suicidality from those MDD without suicidality. Using Optuna optimization, the analysis achieved an area under the curve (AUC) of 0.8117, indicating very good discriminative ability. The diagonal dashed line represents random classification (AUC = 0.5). TPR: True Positive Rate; FPR: False Positive Rate.

**Table 5.**
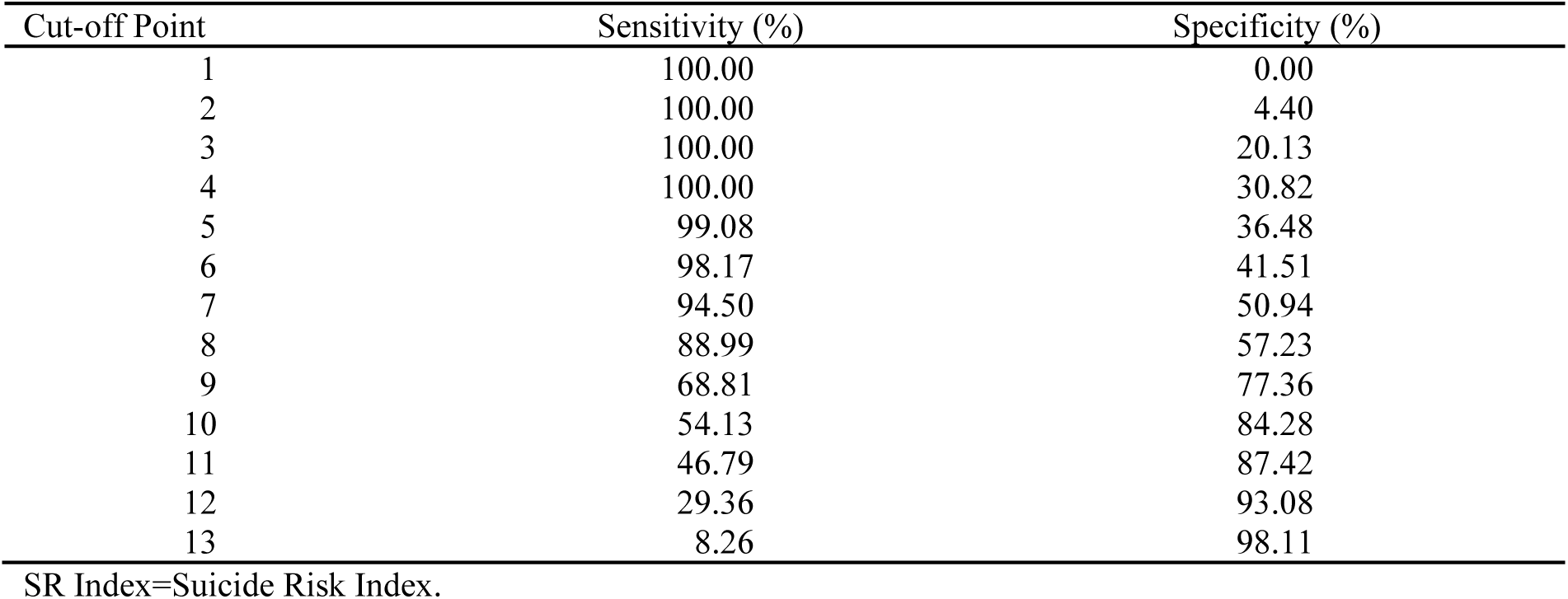
Sensitivity and Specificity Across Different Cut-off Points for Optimal SR Index.

## 4. Discussion

This study introduces an innovative EEG- and ML-based CADx system, approved by the TFDA, for detecting suicidal ideation in MDD patients. A novel SR Index was developed by integrating PHQ-9 and SEA scores, demonstrating excellent discriminative performance (AUC = 0.8117) with significant differences between MDD with suicidality and MDD without suicidality groups (*p* = 2.63×10^-18^). Using an Optuna-optimized cutoff of 8, the SR Index achieved high sensitivity (88.99%) and moderate specificity (57.23%), offering a practical and effective tool for clinical and community settings to identify individuals at high suicide risk.

Extensive research has consistently demonstrated that suicidal ideation and previous suicide attempts are paramount predictors of suicide risk [20]. Suicidal ideation represents the critical inception point of suicidal thoughts and behaviors, serving as a crucial predictor of suicide attempts [21]. Moreover, individuals who have made serious suicide attempts have reported experiencing suicidal thoughts persisting for weeks to months [22]. Among psychiatric conditions, depression is the most common diagnosis in individuals who die by suicide and strongly predicts future suicidal ideation, attempts, and mortality [23]. Given these associations, our study focused specifically on assessing suicidal ideation within the context of depression. Compared with conventional clinical interviews, structured assessments of suicidal ideation can significantly improve the identification of high-risk individuals [24]. Therefore, we used the HDRS-17 suicide item to differentiate between MDD with suicidality and MDD without suicidality, though no standardized protocol exists to assess suicidal ideation to date [24].

Recent advances in neurotechnology based on neurophysiological changes have introduced the possibility of using objective biomarkers to complement clinical assessments. EEG is potential biomarker for depression within the NIMH Research Domain Criteria(RDoC) framework [6, 25]. Resting-state EEG has shown particular promise in identifying neurophysiological alterations associated with MDD and suicide. Studies have demonstrated that depressed individuals exhibit greater right frontal lobe activation compared to healthy controls [26], with specific EEG features such as BP and alpha asymmetry extensively studied as biomarkers [27–31]. MDD patients characteristically show increased functional connectivity in the frontal region for alpha waves and decreased functional connectivity in the parietal-occipital regions [27], and depressed patients exhibit significantly reduced delta wave connectivity during resting state [32]. Furthermore, the neurophysiology of suicide presents distinct patterns. In individuals with suicidal ideation, increased theta wave power at the Cz electrode and fronto-central regions strongly correlates with suicidal thoughts [28], while women with MDD with suicidality exhibit lower beta and gamma wave activity in frontal regions and increased alpha power in posterior regions [29]. Individuals with suicidal ideation showed notably elevated high-gamma activity in frontal, central, and temporal regions, including F4, Fz, C4, Cz, O2, F8, T5, and T6 [30]. Lower frontal alpha asymmetry (FAA), characterized by reduced alpha power in the left frontal region, was observed in MDD with suicidality patients compared to those without suicidal ideation, suggesting that increased right cortical activation associates with higher levels of negative affect[31]. MDD patients experiencing worsening suicidal ideation during antidepressant treatment exhibit significant, albeit brief, decreases in quantitative electroencephalographic cordance in the midline-and-right-frontal region [33]. Regarding suicide attempts, elevated delta wave power in frontal and central regions and abnormal alpha network connectivity show stronger associations with attempts than ideation [28, 34]. Additionally, reduced beta and gamma wave power in the frontal and right temporal regions has been linked to suicide attempts in MDD patients [28, 29].

EEG-based methods have shown potential for classifying suicidal ideation, but previous studies often suffer from limited sample sizes and methodological constraints. Prior research reported that beta wave activity recorded during the reading of potential suicide notes achieved a classification accuracy exceeding 70% for detecting suicidal ideation but with only 16 participants [35]. Similarly, another study demonstrated that high alpha band network indices could distinguish between suicide attempts and ideations with a classification accuracy of 73.39%, sensitivity of 76.36%, and specificity of 70.37% [20]. Yet another approach, combining cognitive tasks and resting-state EEG in an independent sample of 12 participants with suicidality and 23 healthy controls, yielded sensitivity and specificity values of only 50% and 61%, respectively [36]. In contrast, our study’s larger sample of 268 participants achieved superior results, with the SR Index showing an AUC of 0.8117, indicating very good accuracy [18]. With an 8-point cutoff, the SR Index effectively distinguished between MDD with suicidality and MDD without suicidality with a sensitivity of 88.99%, and specificity of 57.23%. The balanced accuracy reached 73.11%, demonstrating the SR Index’s effectiveness in identifying the majority of patients with suicidality while maintaining control over false positives.

Although higher sensitivity can reduce the chances of missing individuals at risk of suicide, it is important to note that excessively high sensitivity (e.g., setting the cutoff between 1 and 4 points, yielding sensitivity of 100%) may lower specificity, leading to an increased rate of false positives (i.e., patients without suicidality incorrectly identified as high-risk). This could result in wasted clinical resources and cause unnecessary anxiety for patients. Conversely, excessively high specificity (e.g., setting the cutoff between 12 and 13 points, yielding specificity between 93.08% and 98.11%) could reduce sensitivity, leading to missed diagnoses among genuinely at-risk patients. Thus, balancing sensitivity and specificity is challenging and should be adjusted according to specific contexts. In screening applications, priority is typically given to improving sensitivity to minimize false negatives. Therefore, setting the SR Index cutoff at 8 points provides an optimal balance between sensitivity and specificity, which is particularly important in clinical screening settings where early detection and intervention for high-risk patients are crucial, such as in primary care or outpatient clinics.

Despite the promising discriminative ability of the SR Index, certain limitations warrant consideration. First, the single-center design with a limited sample size may restrict result generalizability. Second, the heterogeneous nature of both depression symptoms (including various features like anxious distress and melancholic features) and suicidal ideation (ranging from passive to active) could affect external validity. Third, moderate specificity may increase false positives, suggesting the need for multi-stage screening strategies incorporating additional risk factors such as life stress events, family history, or highly specific physiological markers. Lastly, this study focused primarily on screening for suicidal ideation rather than comprehensively predicting suicide attempts or behaviors. Future research should include larger, multi-center samples, consider symptom heterogeneity, implement refined screening protocols, and incorporate data on suicide attempts to better understand the SR Index’s broader applicability.

## 5. Conclusions

The SR Index, integrating PHQ-9 and SEA Index, improves the detection of suicidality in MDD patients with high sensitivity and moderate specificity. Its reliability, ease of use, and efficiency make it a practical tool for schools, communities, and clinical settings. As a complement to structured scales or diagnostic interviews, it supports effective suicide risk screening. Future research should focus on validating its utility in larger and diverse populations to enhance its application in suicide prevention.

## List of Abbreviations

HDRS-17: 17-item Hamilton Depression Rating Scale
BP: Band Power
CADx: Computer-aided diagnosis
EEG: Electroencephalogram
HFD: Higuchi Fractal Dimension
KFD: Katz Fractal Dimension
MDD: Major Depressive Disorder
ML: Machine Learning
PHQ-9: Patient Health Questionnaire-9
TFDA: Taiwan Food and Drug Administration
TSGH: Tri-Service General Hospital

## Acknowledgements

We gratefully acknowledge all attending physicians in the Department of Psychiatry at Tri-Service General Hospital for engaging discussions about the topics in this paper. Preliminary results was presented at the 63rd Annual Meeting of the Taiwanese Society of Psychiatry (2024) in Taipei, Taiwan.

## Author contributions

C.-C. Huang and H.-A. Chang conceptualized the study. C.-L. Hsu, Y.-G. Wang, T.-Y. Chen, C.-Y. Chen, and T.-C. Yeh provided clinical expertise and study-related insights. T.-H. Huang, F.-Y. Yu, and Y.-H. Liu provided software and statistical support. C.-C. Huang wrote the initial manuscript draft. C.-C. Chang and H.-A. Chang conducted critical revision of the manuscript and served as corresponding authors. All authors reviewed and approved the final version of the manuscript.

## Funding

This study was supported in part by grants from Tri-Service General Hospital (TSGH-D-113197) and Ministry of National Defense (MND-MAB-D-111138 and MND-MAB-D-112164). The funders had no role in the design of the study; in the collection, analyses, or interpretation of data; in the writing of the manuscript, or in the decision to publish the results.

## Data availability

The datasets used and/or analyzed in this study are available from the corresponding author upon reasonable request.

## Declarations

### Ethics approval and consent to participate

This study was conducted in accordance with the ethical principles outlined in the Declaration of Helsinki and was approved by the Institutional Review Board (IRB) of Tri-Service General Hospital (IRB# A202305083). Since the study involved a retrospective review of data, the requirement for informed consent was waived. All methods were carried out in compliance with relevant guidelines and regulations.

Clinical trial number: not applicable.

### Consent for publication

Not applicable.

### Competing interests

All authors declare no financial or personal relationships with individuals or organizations that could unduly influence work.

## Notes

### Competing Interest Statement

The authors have declared no competing interest.

### Author Declarations

Institutional Review Board (IRB) of Tri-Service General Hospital

## References

1. American Psychiatric Association D, American Psychiatric Association D. Diagnostic and statistical manual of mental disorders: DSM-5, vol. 5. American psychiatric association Washington, DC; 2013.

2. Orsolini L, Latini R, Pompili M, Serafini G, Volpe U, Vellante F, Fornaro M, Valchera A, Tomasetti C, Fraticelli S et al. Understanding the Complex of Suicide in Depression: from Research to Clinics. Psychiatry Investig. 2020; 17(3):207–21.

3. Arias-de la Torre J, Vilagut G, Ronaldson A, Serrano-Blanco A, Martín V, Peters M, Valderas JM, Dregan A, Alonso J. Prevalence and variability of current depressive disorder in 27 European countries: a population-based study. The Lancet Public Health. 2021; 6(10):e729–e738.

4. Wu CT, Huang HC, Huang S, Chen IM, Liao SC, Chen CK, Lin C, Lee SH, Chen MH, Tsai CF et al. Resting-State EEG Signal for Major Depressive Disorder Detection: A Systematic Validation on a Large and Diverse Dataset. Biosensors (Basel). 2021; 11(12).

5. Global, regional, and national incidence, prevalence, and years lived with disability for 354 diseases and injuries for 195 countries and territories, 1990-2017: a systematic analysis for the Global Burden of Disease Study 2017. Lancet. 2018; 392(10159):1789–858.

6. de Aguiar Neto FS, Rosa JLG. Depression biomarkers using non-invasive EEG: A review. Neurosci Biobehav Rev. 2019; 105:83–93.

7. Organization WH. Suicide worldwide in 2019: global health estimates. 2021.

8. Fazel S, Runeson B. Suicide. N Engl J Med. 2020; 382(3):266–74.

9. Goldberg D. The heterogeneity of “major depression“. World Psychiatry. 2011; 10(3):226–28.

10. Kroenke K, Spitzer RL, Williams JB. The PHQ-9: validity of a brief depression severity measure. J Gen Intern Med. 2001; 16(9):606–13.

11. Runeson B, Odeberg J, Pettersson A, Edbom T, Jildevik Adamsson I, Waern M. Instruments for the assessment of suicide risk: A systematic review evaluating the certainty of the evidence. PLoS One. 2017; 12(7):e0180292.

12. Su C, Aseltine R, Doshi R, Chen K, Rogers SC, Wang F. Machine learning for suicide risk prediction in children and adolescents with electronic health records. Translational psychiatry. 2020; 10(1):413.

13. Mehta S, Downar J, Mulsant BH, Voineskos D, Daskalakis ZJ, Weissman CR, Vila-Rodriguez F, Blumberger DM. Effect of high frequency versus theta-burst repetitive transcranial magnetic stimulation on suicidality in patients with treatment-resistant depression. Acta Psychiatr Scand. 2022; 145(5):529–538.

14. Higuchi T. Approach to an irregular time series on the basis of the fractal theory. Physica D: Nonlinear Phenomena. 1988; 31(2):277–83.

15. Katz MJ. Fractals and the analysis of waveforms. Comput Biol Med. 1988; 18(3):145–56.

16. Bermingham ML, Pong-Wong R, Spiliopoulou A, Hayward C, Rudan I, Campbell H, Wright AF, Wilson JF, Agakov F, Navarro P et al. Application of high-dimensional feature selection: evaluation for genomic prediction in man. Sci Rep. 2015; 5:10312.

17. Cortes C. Support-Vector Networks. Machine Learning. 1995.

18. Okeh U, Okoro C. Evaluating measures of indicators of diagnostic test performance: fundamental meanings and formulars. J Biom Biostat. 2012; 3(1):2.

19. Akiba T, Sano S, Yanase T, Ohta T, Koyama M. Optuna: A next-generation hyperparameter optimization framework. In: Proceedings of the 25th ACM SIGKDD international conference on knowledge discovery & data mining. 2019.

20. Posner K, Brown GK, Stanley B, Brent DA, Yershova KV, Oquendo MA, Currier GW, Melvin GA, Greenhill L, Shen S et al. The Columbia-Suicide Severity Rating Scale: initial validity and internal consistency findings from three multisite studies with adolescents and adults. Am J Psychiatry. 2011; 168(12):1266–77.

21. Klonsky ED, May AM, Saffer BY. Suicide, Suicide Attempts, and Suicidal Ideation. Annu Rev Clin Psychol. 2016; 12:307–30.

22. Anestis MD, Soberay KA, Gutierrez PM, Hernández TD, Joiner TE. Reconsidering the link between impulsivity and suicidal behavior. Pers Soc Psychol Rev. 2014; 18(4):366–86.

23. Ribeiro JD, Huang X, Fox KR, Franklin JC. Depression and hopelessness as risk factors for suicide ideation, attempts and death: meta-analysis of longitudinal studies. Br J Psychiatry. 2018; 212(5):279–86.

24. Bongiovi-Garcia ME, Merville J, Almeida MG, Burke A, Ellis S, Stanley BH, Posner K, Mann JJ, Oquendo MA. Comparison of clinical and research assessments of diagnosis, suicide attempt history and suicidal ideation in major depression. J Affect Disord. 2009; 115(1-2):183–8.

25. Insel T, Cuthbert B, Garvey M, Heinssen R, Pine DS, Quinn K, Sanislow C, Wang P. Research domain criteria (RDoC): toward a new classification framework for research on mental disorders. Am J Psychiatry. 2010; 167(7):748–51.

26. Schaffer CE, Davidson RJ, Saron C. Frontal and parietal electroencephalogram asymmetry in depressed and nondepressed subjects. Biol Psychiatry. 1983; 18(7):753–62.

27. Miljevic A, Bailey NW, Murphy OW, Perera MPN, Fitzgerald PB. Alterations in EEG functional connectivity in individuals with depression: A systematic review. J Affect Disord. 2023; 328:287–302.

28. Amico F, Frye RE, Shannon S, Rondeau S. Resting State EEG Correlates of Suicide Ideation and Suicide Attempt. J Pers Med. 2023; 13(6).

29. Benschop L, Baeken C, Vanderhasselt MA, Van de Steen F, Van Heeringen K, Arns M. Electroencephalogram Resting State Frequency Power Characteristics of Suicidal Behavior in Female Patients With Major Depressive Disorder. J Clin Psychiatry. 2019; 80(6).

30. Arikan MK, Gunver MG, Tarhan N, Metin B. High-Gamma: A biological marker for suicide attempt in patients with depression. J Affect Disord. 2019; 254:1–6.

31. Roh SC, Kim JS, Kim S, Kim Y, Lee SH. Frontal Alpha Asymmetry Moderated by Suicidal Ideation in Patients with Major Depressive Disorder: A Comparison with Healthy Individuals. Clin Psychopharmacol Neurosci. 2020; 18(1):58–66.

32. Huang SS, Yu YH, Chen HH, Hung CC, Wang YT, Chang CH, Peng SJ, Kuo PH. Functional connectivity analysis on electroencephalography signals reveals potential biomarkers for treatment response in major depression. BMC Psychiatry. 2023; 23(1):554.

33. Hunter AM, Leuchter AF, Cook IA, Abrams M. Brain functional changes (QEEG cordance) and worsening suicidal ideation and mood symptoms during antidepressant treatment. Acta Psychiatr Scand. 2010; 122(6):461–69.

34. Kim S, Jang KI, Lee HS, Shim SH, Kim JS. Differentiation between suicide attempt and suicidal ideation in patients with major depressive disorder using cortical functional network. Prog Neuropsychopharmacol Biol Psychiatry. 2024; 132:110965.

35. Prasad DK, Liu S, Chen S-HA, Quek C. Sentiment analysis using EEG activities for suicidology. Expert Systems with Applications. 2018; 103:206–17.

36. Nan J, Grennan G, Ravichandran S, Ramanathan D, Mishra J. Neural activity during inhibitory control predicts suicidal ideation with machine learning. NPP Digit Psychiatry Neurosci. 2024; 2(1):10.

